# Disturbed sleep after lung transplantation is associated with worse patient-reported outcomes and chronic lung allograft dysfunction

**DOI:** 10.1101/2023.10.12.23296973

**Authors:** Aric A. Prather, Ying Gao, Legna Betancourt, Rose C Kordahl, Anya Sriram, Chiung-Yu Huang, Steven R Hays, Jasleen Kukreja, Daniel R. Calabrese, Aida Venado, Bhavya Kapse, John R Greenland, Jonathan P Singer

## Abstract

Many lung transplant recipients fail to derive the expected improvements in functioning, HRQL, or long-term survival. Sleep may represent an important, albeit rarely examined, factor influencing lung transplant outcomes.

Within a larger cohort study, 141 lung transplant recipients completed the Medical Outcomes Study (MOS) Sleep Scale along with a broader survey of patient-reported outcome (PRO) measures and frailty assessment. MOS Sleep yields the Sleep Problems Index (SPI); we also derived an insomnia-specific subscale. Potential perioperative predictors of disturbed sleep and time to chronic lung allograft dysfunction (CLAD) and death were derived from medical records. We investigated associations between perioperative predictors on SPI and Insomnia and associations between SPI and Insomnia on PROs and frailty by linear regressions, adjusting for age, sex, and lung function. We evaluated the associations between SPI and Insomnia on time to CLAD and death using Cox models, adjusting for age, sex, and transplant indication.

Post-transplant hospital length of stay >30 days was associated with worse sleep by SPI and insomnia (SPI: p=0.01; Insomnia p=0.02). Worse sleep by SPI and insomnia was associated with worse depression, cognitive function, HRQL, physical disability, health utilities, and Fried Frailty Phenotype frailty (all p<0.01). Those in the worst quartile of SPI and insomnia exhibited increased risk of CLAD (HR 2.18; 95%CI: 1.22—3.89; p=0.01 for SPI and HR 1.96; 95%CI 1.09—3.53; p=0.03 for insomnia). Worsening in SPI but not insomnia was also associated with mortality (HR: 1.29; 95%CI: 1.05—1.58; p=0.01).

Poor sleep after lung transplant may be a novel predictor of patient reported outcomes, frailty, CLAD, and death with potentially important screening and treatment implications.

## Introduction

Lung transplantation aims to extend survival, relieve disability, and improve health-related quality of life (HRQL).^1,2^ Although many do well, perioperative complications are increasing, 20-30% of survivors do not report substantive improvements in physical functioning or HRQL, and one in three die within the first three post-operative years,.^1,3–8^ In response to these frequent and increasing barriers to transplant success, identifying modifiable factors that could support transplant recovery and improve HRQL is needed. Sleep may be one potential behavioral pathway, although its impacts on outcomes after lung transplantation are not well characterized.

Sleep plays a fundamental role in physical health, well-being, and recovery from illness. The potential for sleep disturbances after lung transplant is high, particularly during the dynamic peri- and early post-operative period that is increasingly complicated by Primary Graft Dysfunction (PGD; a form of acute post-transplant lung injury), delirium, prolonged hospital stays, post-traumatic stress, readmissions, and intense immunosuppression.^6,9–13^ Disturbed sleep has the potential to significantly impair transplant success. Outside of the lung transplant context, disturbed sleep is associated with impaired cognition, mood dysregulation (particularly depression), risk of frailty, and immune dysfunction.^14–22^ In addition to being important outcomes to lung transplant recipients themselves, depression, frailty and immune dysfunction have previously been shown to also predict disability, chronic lung graft dysfunction (CLAD), and mortality after transplant.^23–27^

To date, a relatively limited number of studies have examined sleep after lung transplantation and even fewer have examined the impact of disturbed sleep on post-transplant outcomes.^28^ Depending on the sleep measure used, between 30-74% of lung transplant recipients reported disturbed sleep; when present, disturbed sleep was associated with depression and anxiety.^28^

To address these gaps, in this single-center cohort study we sought to examine potential perioperative predictors of disturbed sleep after lung transplant and whether disturbed sleep is associated with disability, HRQL, and frailty. We additionally sought to test whether disturbed sleep was associated with risk of CLAD onset and mortality.

## Methods

### Study Design

We performed this study among a subset of participants of the UCSF “Breathe Again” cohort. “Breathe Again” is a single-center, longitudinal, repeated measure prospective cohort study of 259 adults who underwent first-time lung transplantation between 2010 and 2017.^29^ Between November 2013 and August 2016, 200 participants completed a supplemental pilot patient-reported outcome survey battery once during one of their routine study visits. This pilot survey ultimately yielded the Lung Transplant Quality of Life (LT-QOL) measure details of the pilot survey composition and administration are detailed in reporting of LT-QOL development.^30^ Of the 200 participants who completed the pilot survey, 141 completed a version that included the Medical Outcomes Study Sleep Scale (MOS-Sleep)^31^; it is this group of 141 that forms the group analyzed herein. Our study was approved by the UCSF Institutional Review Board.

### Measures of Sleep

The MOS-Sleep is a 6-item validated measure of sleep disturbance that is scored as the Sleep Problems Index (SPI; range: 0 to 100; higher scores denote worse sleep).^31^ To further understand the specific impact of insomnia, we also explored an insomnia-specific subscale based on the face validity of two of the six MOS-Sleep items. These insomnia-focused items query how often respondents had trouble falling asleep and how often respondents awakened during sleep time and then had trouble falling asleep again.

### Predictor variables of interest

Based on sleep literature from other medical and surgical patient populations, we considered age, sex, transplant indication, and BMI as well as intensive care unit (ICU) delirium, prolonged hospital length of stay, and, for lung transplant, severe Primary Graft Dysfunction (PGD) as potential demographic and perioperative causes of disturbed sleep after transplant. ICU delirium was defined as ever being delirious in the ICU after lung transplant surgery by medical record review of nursing-administered Confusion Assessment Method for the ICU (CAM-ICU)^32^ screens once per shift. Severe PGD was defined as Grade 3 PGD on post-operative day 2 or 3 per International Society of Heart and Lung Transplantation (ISHLT) consensus.^33^ Prolonged hospital length of stay was defined as post-transplant length of stay lasting 30 or more days after transplant.^6^

### Outcome Variables of Interest

All participants in *Breathe Again* completed a complex study battery of patient reported battery (PROs) and frailty before and repeatedly up to 36 months after transplant. For this analysis, we selected the survey responses and frailty assessments after transplant that were collected concurrent with the pilot survey.

### Patient reported outcomes

The survey measures included instruments to assess functioning/disability, depressive symptoms, generic and respiratory-specific HRQL, and health utilities. In addition to the Sleep Scale, the pilot survey also included the original Medical Outcomes Study Cognitive Functioning Scale (MOS-Cog)^31^ that has been rescaled and adapted to become the Lung Transplant Quality of Life Cognitive Limitations subscale (LTQOL-Cog).^30^ Functioning/disability was assessed by the Lung Transplant Valued Life Activities Scale (LT-VLA; 15 items; range 0-3; Minimally important difference [MID]: 0.3; higher scores denote worse disability. Depressive symptoms were quantified by the Geriatrics Depression Scale-15 (GDS; 15 items; range 0-15; MID: 1.65; higher scores denote worse depressive symptoms). Generic HRQL was evaluated by the RAND Medical Outcomes Study Short Form-36 Physical and Mental Composite Summary scales (SF-36 PCS and MCS; 36 items; range 0-100; MID 5; lower scores denote worse HRQL). Respiratory-specific HRQL was assessed with the Airways Questionnaire 20-Revised (AQ20-R; 20 items; range 0-20; MID 1.75; higher scores denote worse HRQL). Health utility was assessed by the EuroQol 5D (EQ5D; 5 items; range −0.11 – 1.0; MID 0.06; higher scores denote better health utility). Finally self-reported cognitive functioning was assessed by the LTQOL-Cog (6 items; range 1-5; MID: 0.47; higher scores denote worse cognitive functioning).

### Frailty

We assessed frailty by two well-validated frailty measures that emphasize physical functioning. The Short Physical Performance Battery (SPPB) is a 3-component battery of lower extremity performance measures that includes gait speed, chair stands, and balance ^34,35^. Each measure is scored from 0–4 with an aggregate score ranging from 0–12. Lower SPPB scores reflect increased frailty. The Fried Frailty Phenotype (FFP) is an aggregate score of five constructs: shrinking, exhaustion, low physical activity, slowness, and weakness.^36^ The FFP ranges from 0–5 with higher scores reflecting increased frailty.

### *Chronic Lung Allograft Dysfunction* (CLAD)

CLAD was defined as 20% decline in forced expiratory volume in 1 second (FEV1) from post-transplant baseline that persisted for at least 3 months.^37^ All pulmonary lung function after transplant were abstracted from medical records. Time to CLAD was calculated as the number of days from the date of lung transplantation until the first sustained 20% drop in FEV1.

### Mortality

Dates of death were obtained through the Social Security Master Death File. Survival time was calculated as the number of days from the date of lung transplantation until date of death.

### Other measurements

Demographic and clinical variables were abstracted from medical records. Variables included age, gender, race/ethnicity, diagnostic indication for transplant, transplant type (single versus bilateral versus heart-lung) and all measures of lung function (forced expiratory volume in one second [FEV1; liters] and forced vital capacity [FVC; liters] after transplant.

### Analytic Approach

We used the overall SPI and two insomnia-focused items to test the association between both disturbed sleep and insomnia on our outcomes of interest. The SPI and the insomnia subscale do not have established thresholds to define disturbed sleep or insomnia. Conceptually, *any* impairment in sleep could plausibly be associated with how people feel and function. Therefore, we analyzed the SPI and insomnia subscale on a continuous scale (per minimally important difference [MID] worsening) and as binary variables comparing the worst quartile to the rest. Some scales, such as the SF36 and EQ5D, have established anchor-based MIDs. For many PROs, including the MOS-Sleep, anchor-based MIDs are not available. In these cases, distribution-based methods are employed with one-half the observed standard deviation being the most common.^38^ For this analysis, we defined SPI and insomnia MIDs by one-half the observed standard deviation, as we have done previously.^29^

To investigate the associations between demographic, genotypic, and post-operative ICU delirium, PGD, and prolonged hospital LOS variables on SPI and Insomnia as continuous and as binary outcomes, we used linear and logistic regression, respectively, adjusting for age, sex, and pre-operative lung function. To investigate the association between SPI and Insomnia as continuous and binary exposure variables on PROs and frailty we used linear regression, adjusting for age, sex, transplant indication and post-operative lung function. We fit Cox proportional hazards models to evaluate the associations between SPI and Insomnia with time to CLAD and time to death adjusted for age, sex, and transplant indication. The proportional hazards assumption was tested using Schoenfeld residuals. We used Kaplan Meier methods to visualize the relationship association between the SPI and insomnia defined categorically with CLAD and death. We used the Survival Area Plot method to visualize the unadjusted association between the SPI and insomnia as continuous variables with CLAD and death.^39^

Analyses were performed using SAS (version 9.4, SAS Institute), and R (version 4.3.1, R Foundation).

## Results

During the Breathe Again study period, of the 392 participants enrolled, 259 underwent lung transplant. Of these, 141 completed the pilot survey that included MOS-Sleep and formed our study cohort (Table 1 and Figure 1). These 141 participants were 43% female with a mean age of 58 years (standard deviation [SD] ± 13); 75% were non-Hispanic white. Most participants underwent transplant for pulmonary fibrosis (72%) followed by non-suppurative obstructive lung diseases (15%). Participants completed the study battery used in these analyses at a median of 1.5 years after transplant (Interquartile range [IQR]: 0.6, 2.4). Over the study period, 52 (37%) developed CLAD and 20 (14%) died.

**Figure 1.**
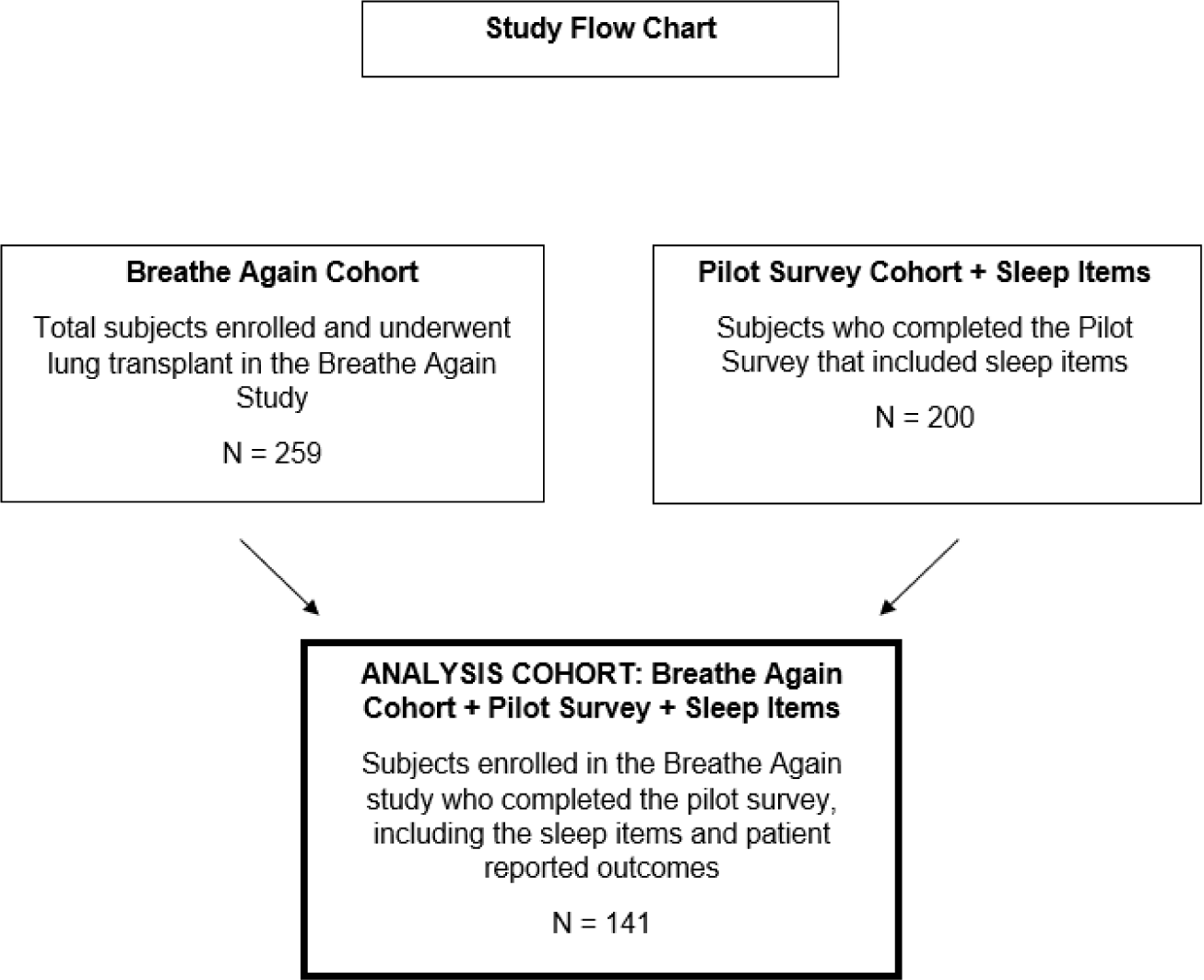
Study Flow

**Table 1.**
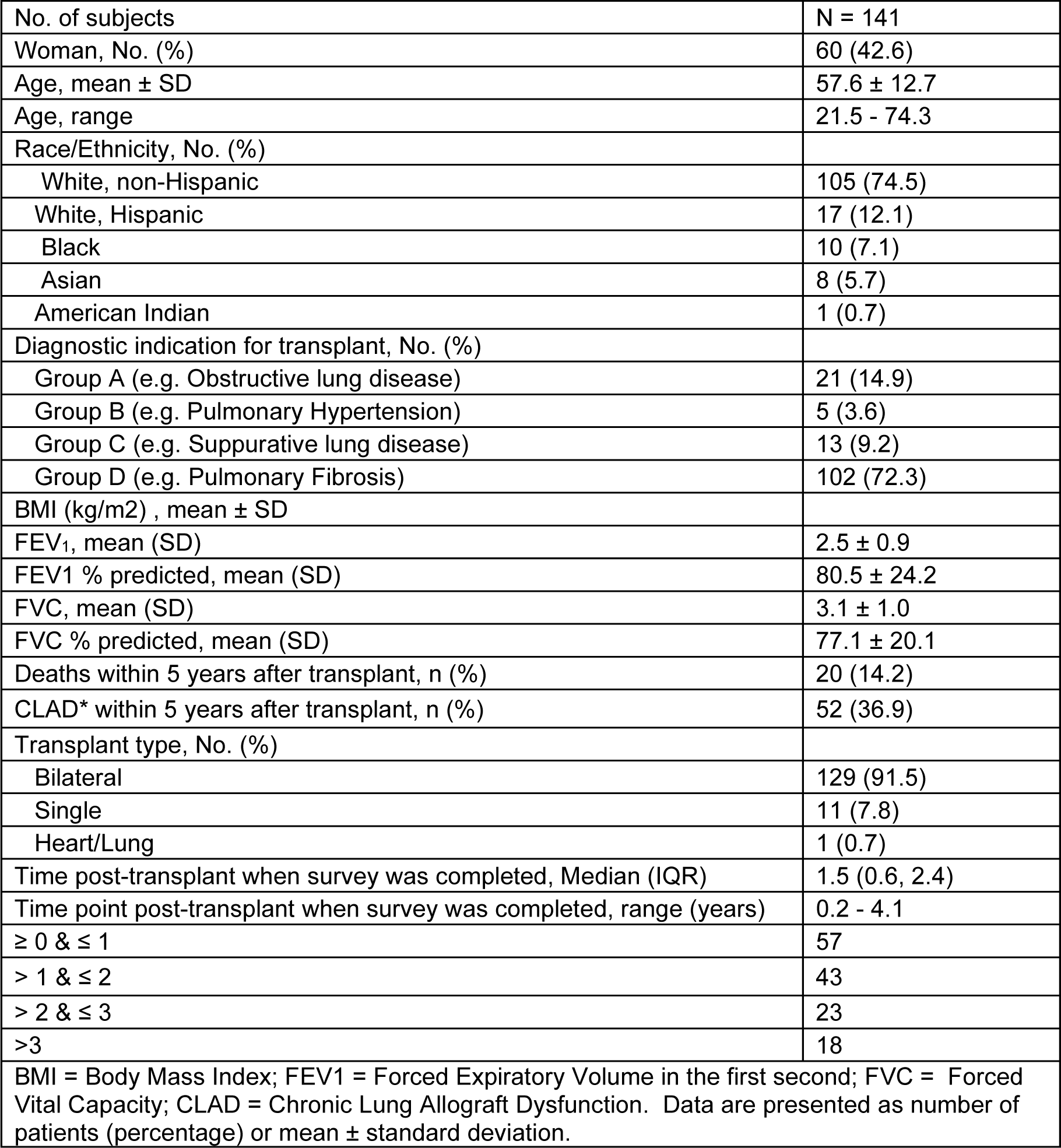
Participant characteristics.

|  |  |
| --- | --- |
| No. of subjects | N = 141 |
| Woman, No. (%) | 60 (42.6) |
| Age, mean ± SD | 57.6 ± 12.7 |
| Age, range | 21.5 - 74.3 |
| Race/Ethnicity, No. (%) |  |
| White, non-Hispanic | 105 (74.5) |
| White, Hispanic | 17 (12.1) |
| Black | 10 (7.1) |
| Asian | 8 (5.7) |
| American Indian | 1 (0.7) |
| Diagnostic indication for transplant, No. (%) |  |
| Group A (e.g. Obstructive lung disease) | 21 (14.9) |
| Group B (e.g. Pulmonary Hypertension) | 5 (3.6) |
| Group C (e.g. Suppurative lung disease) | 13 (9.2) |
| Group D (e.g. Pulmonary Fibrosis) | 102 (72.3) |
| BMI (kg/m<sup>2</sup>) , mean ± SD | |
| FEV<sub>1</sub>, mean (SD) | 2.5 ± 0.9 |
| FEV1 % predicted, mean (SD) | 80.5 ± 24.2 |
| FVC, mean (SD) | 3.1 ± 1.0 |
| FVC % predicted, mean (SD) | 77.1 ± 20.1 |
| Deaths within 5 years after transplant, n (%) | 20 (14.2) |
| CLAD* within 5 years after transplant, n (%) | 52 (36.9) |
| Transplant type, No. (%) |  |
| Bilateral | 129 (91.5) |
| Single | 11 (7.8) |
| Heart/Lung | 1 (0.7) |
| Time post-transplant when survey was completed, Median (IQR) | 1.5 (0.6, 2.4) |
| Time point post-transplant when survey was completed, range (years) | 0.2 - 4.1 |
| $\geq 0$ & $\leq 1$ | 57 |
| $> 1$ & $\leq 2$ | 43 |
| $> 2$ & $\leq 3$ | 23 |
| $> 3$ | 18 |
| BMI = Body Mass Index; FEV1 = Forced Expiratory Volume in the first second; FVC = Forced Vital Capacity; CLAD = Chronic Lung Allograft Dysfunction. Data are presented as number of patients (percentage) or mean ± standard deviation. | |

Of the predictors tested, prolonged post-transplant hospital length of stay (LOS) was associated with disturbed sleep and insomnia, after adjusting for age, sex, and lung function. Considering SPI and insomnia as continuous measures, prolonged LOS was associated with 12-points worse SPI scores (12.18; 95%CI: 3.38, 20.99; p = 0.01; SPI MID = 8) and 15-points worse insomnia scores (14.56; 95%CI: 2.08, 27.04; p = 0.02; insomnia MID = 12). Prolonged LOS was associated with a 7-fold higher odds of being in the worst quartile of SPI disturbed sleep (adjusted Odds Ratio [aOR] 7.09; 95%CI: 2.10, 23.95; p <0.01). Prolonged LOS, however, was not significantly associated with being in the worst quartile of insomnia (aOR: 1.69; 95%CI: 0.47, 6.04; p = 0.42). Neither PGD nor delirium were significantly associated with disturbed sleep or insomnia (all p >0.05) (Appendix Table 1).

We also found consistent associations between disturbed sleep, either as a continuous variable or dichotomized as worst versus the top three quartiles, and all patient-reported outcomes of interest, after adjusting for age, sex, transplant indication, and allograft function (Table 2). For example, for each MID worsening in the Sleep Problems Index (SPI) participants reported worse depressive symptoms (GDS: 0.63; 95%CI: 0.32, 0.94; p < 0.01; MID = 1.65), generic mental HRQL (SF36 MCS: −2.04; 95%CI: −2.75, −1.33; p <0.01; MID=5), and health utilities (EQ5D: −0.03; 95%CI: −0.04, −0.02; p<0.01; MID=0.06). Similarly, participants in worst quartile of SPI reported substantially worse respiratory-specific HRQL (AQ20-R: 3.53; 95%CI: 1.79, 5.26; p<0.01; MID=1.75), cognitive functioning (LTQOL-Cog: 0.63; 95%CI: 0.31, 0.96; p <0.01; MID=0.47), and generic physical HRQL (SF36-PCS: −6.99; 95%CI: −10.67, −3.30; p <0.01; MID=5). Insomnia symptoms were similarly strongly associated with our PROs of interest after adjusting for prespecified confounders (Table 3).

**Table 2:**
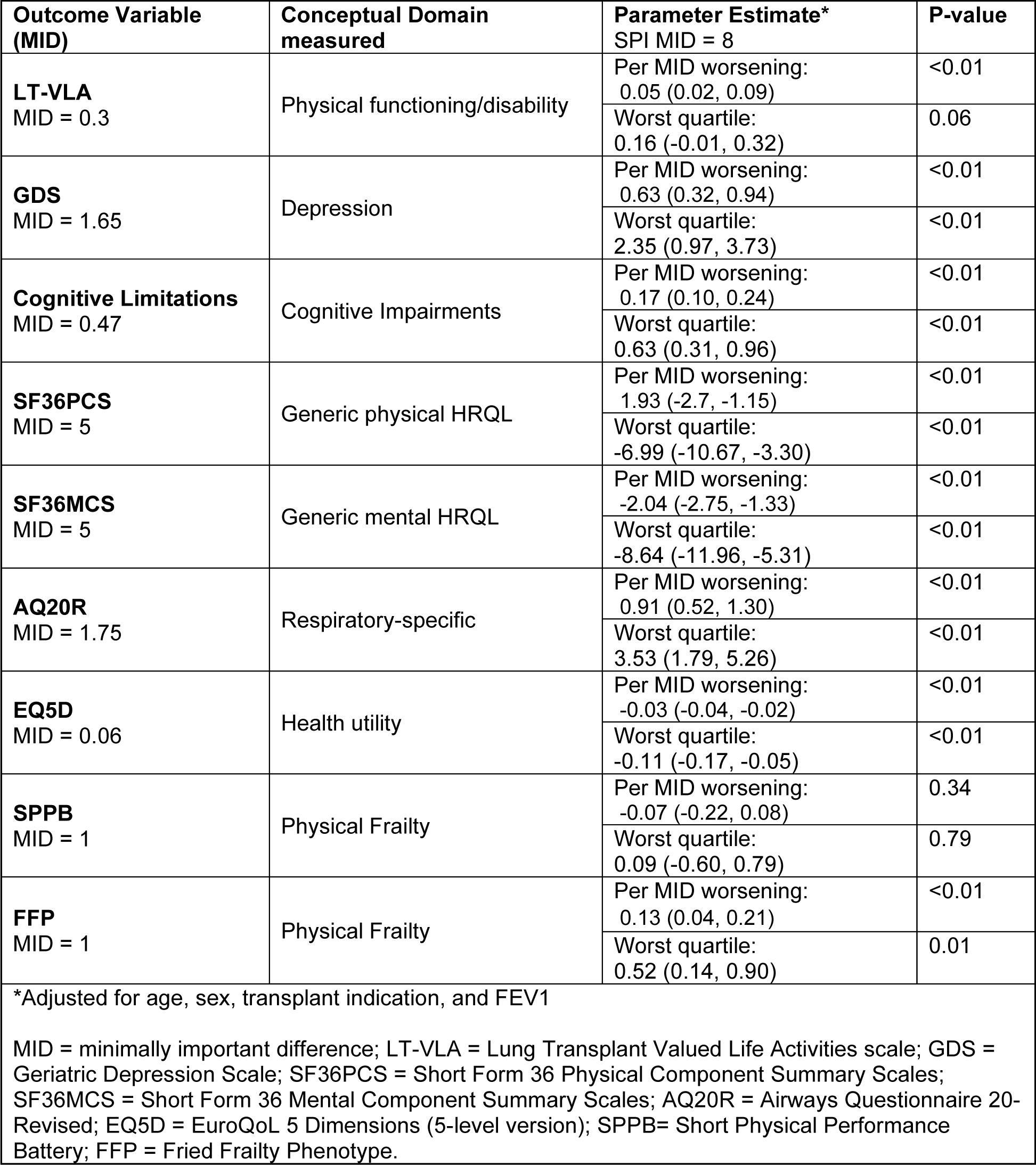
Association between sleep problem index with patient-centered outcomes.

**Table 3:** Association between insomnia and patient-centered outcomes.

| **Outcome Variable (MID)** | **Conceptual Domain measured** | **Parameter Estimate\***<br>Insomnia subscale MID = 12 | **P-value** |
| --- | --- | --- | --- |
| **VLA**<br>MID = 0.3 | Physical functioning/disability | Per MID worsening:<br>0.05 (0.02, 0.09) | <0.01 |
|  |  | Worst quartile:<br>0.22 (0.06, 0.38) | 0.01 |
| **GDS**<br>MID = 1.65 | Depression | Per MID worsening:<br>0.58 (0.25, 0.90) | <0.01 |
|  |  | Worst quartile:<br>2.33 (0.96, 3.70) | <0.01 |
| **Cognitive Limitations**<br>MID = 0.47 | Cognitive impairments | Per MID worsening:<br>0.18 (0.11, 0.25) | <0.01 |
|  |  | Worst quartile:<br>0.71 (0.39, 1.03) | <0.01 |
| **SF36PCS**<br>MID = 5 | Generic physical HRQL | Per MID worsening:<br>-1.76 (-2.59, -0.93) | <0.01 |
|  |  | Worst quartile:<br>-6.57 (-10.28, -2.86) | <0.01 |
| **SF36MCS**<br>MID = 5 | Generic mental HRQL | Per MID worsening:<br>-1.98 (-2.74, -1.22) | <0.01 |
|  |  | Worst quartile:<br>-8.26 (-11.62, -4.90) | <0.01 |
| **AQ20R**<br>MID = 1.75 | Respiratory-specific | Per MID worsening:<br>1.06 (0.67, 1.44) | <0.01 |
|  |  | Worst quartile:<br>4.47 (2.87, 6.07) | <0.01 |
| **EQ5D**<br>MID = 0.06 | Health utility | Per MID worsening:<br>-0.02 (-0.04, -0.01) | <0.01 |
|  |  | Worst quartile:<br>-0.08 (-0.14, -0.02) | <0.01 |
| **SPPB**<br>MID = 1 | Physical Frailty | Per MID worsening:<br>-0.05 (-0.21, 0.11) | 0.54 |
|  |  | Worst quartile:<br>0.13 (-0.57, 0.82) | 0.72 |
| **FFP**<br>MID = 1 | Physical Frailty | Per MID worsening:<br>0.12 (0.03, 0.21) | <0.01 |
|  |  | Worst quartile:<br>0.43 (0.03, 0.82) | 0.03 |

\*Adjusted for age, sex, transplant indication, and FEV1
MID = minimally important difference; LT-VLA = Lung Transplant Valued Life Activities scale; GDS = Geriatric Depression Scale; SF36PCS = Short Form 36 Physical Component Summary Scales; SF36MCS = Short Form 36 Mental Component Summary Scales; AQ20R = Airways Questionnaire 20-Revised; EQ5D = EuroQoL 5 Dimensions (5-level version); SPPB= Short Physical Performance Battery; FFP = Fried Frailty Phenotype.

Poorer sleep and insomnia symptoms were also generally associated with worse physical functioning and frailty by FFP (Tables 2 and 3). For example, each MID worsening in SPI was associated with disability (LT-VLA: 0.05; 95%CI: 0.02, 0.09; p <0.01; MID = 0.3) and frailty (FFP: 0.13; 95%CI: 0.04, 0.21; p <0.01). Each MID worsening in insomnia was also associated with disability (LT-VLA: 0.05; 95%CI: 0.02, 0.09; p <0.01; MID = 0.3) and frailty (FFP: 0.12; 95%CI: 0.03, 0.21; p <0.01). Sleep did not appear to be significantly associated with frailty by SPPB.

Finally, poorer sleep and insomnia also appeared to be associated with the development of CLAD and death after lung transplantation. For example, adjusting for age, sex, and transplant indication, each MID worsening in sleep by SPI was associated with a 1.14-fold increased risk of CLAD (HR 1.14; 95%CI: 1.00, 1.30; p=0.04) and a 1.29-fold increased risk of death (HR: 1.29; 95%CI: 1.05, 1.58; p=0.01). The direction and magnitude of the associations were relatively consistent across analyses, although the strength of associations between binary SPI and insomnia with death after transplant did not reach statistical significance in tests. Results are detailed in Table 4 and in Figures 2 and 3.

**Figure 2.**
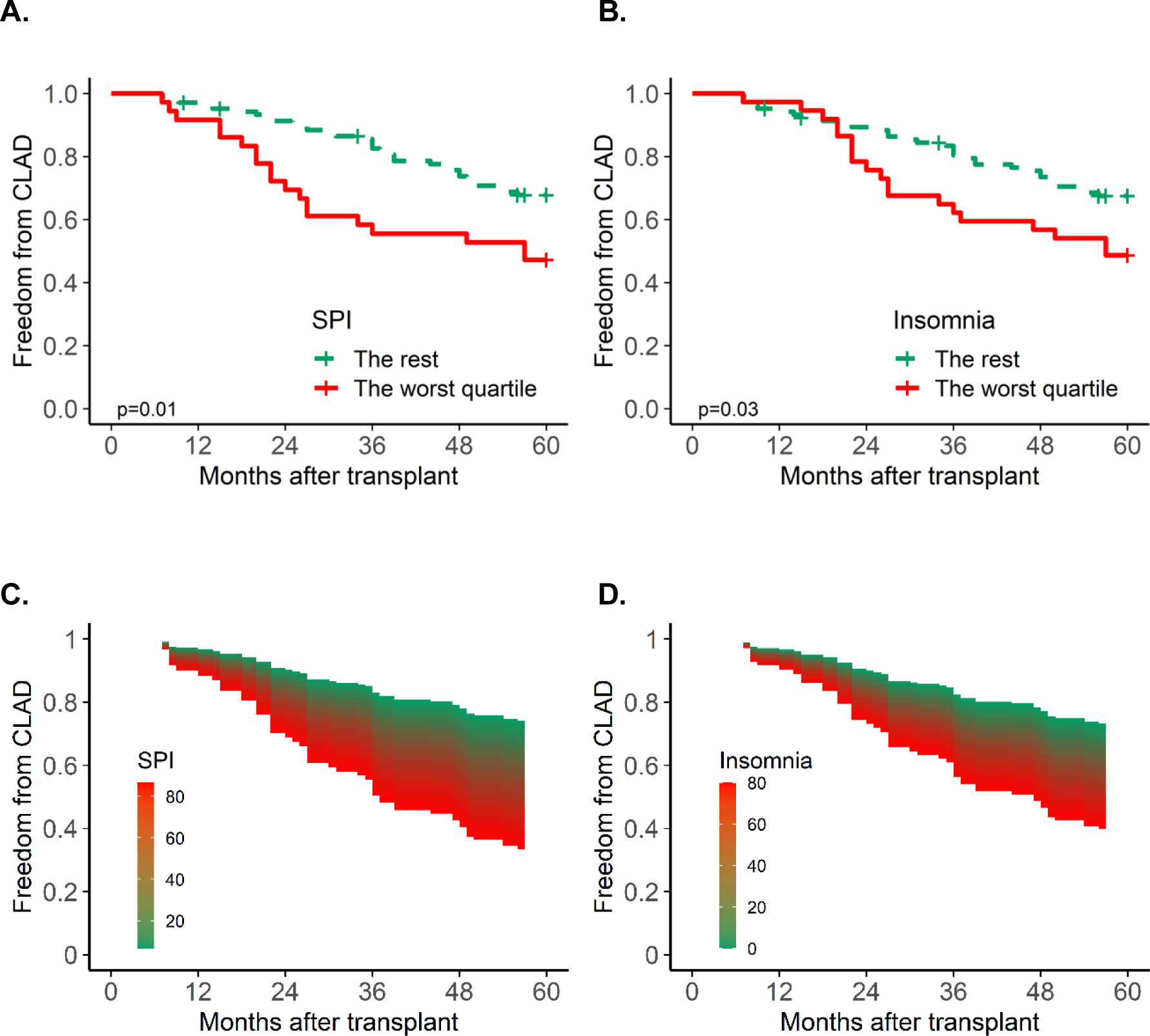
Kaplan Meier estimate of the association of disturbed sleep by MOS-Sleep Problems Index (panel A) and insomnia-specific subscale (panel B) defined as categorical variables on time to CLAD. Survival Area Plot illustrating the association of disturbed sleep by MOS-Sleep Problems Index (panel C) and insomnia-specific subscale (panel D) defined as continuous variables on time to CLAD.

**Figure 3.**
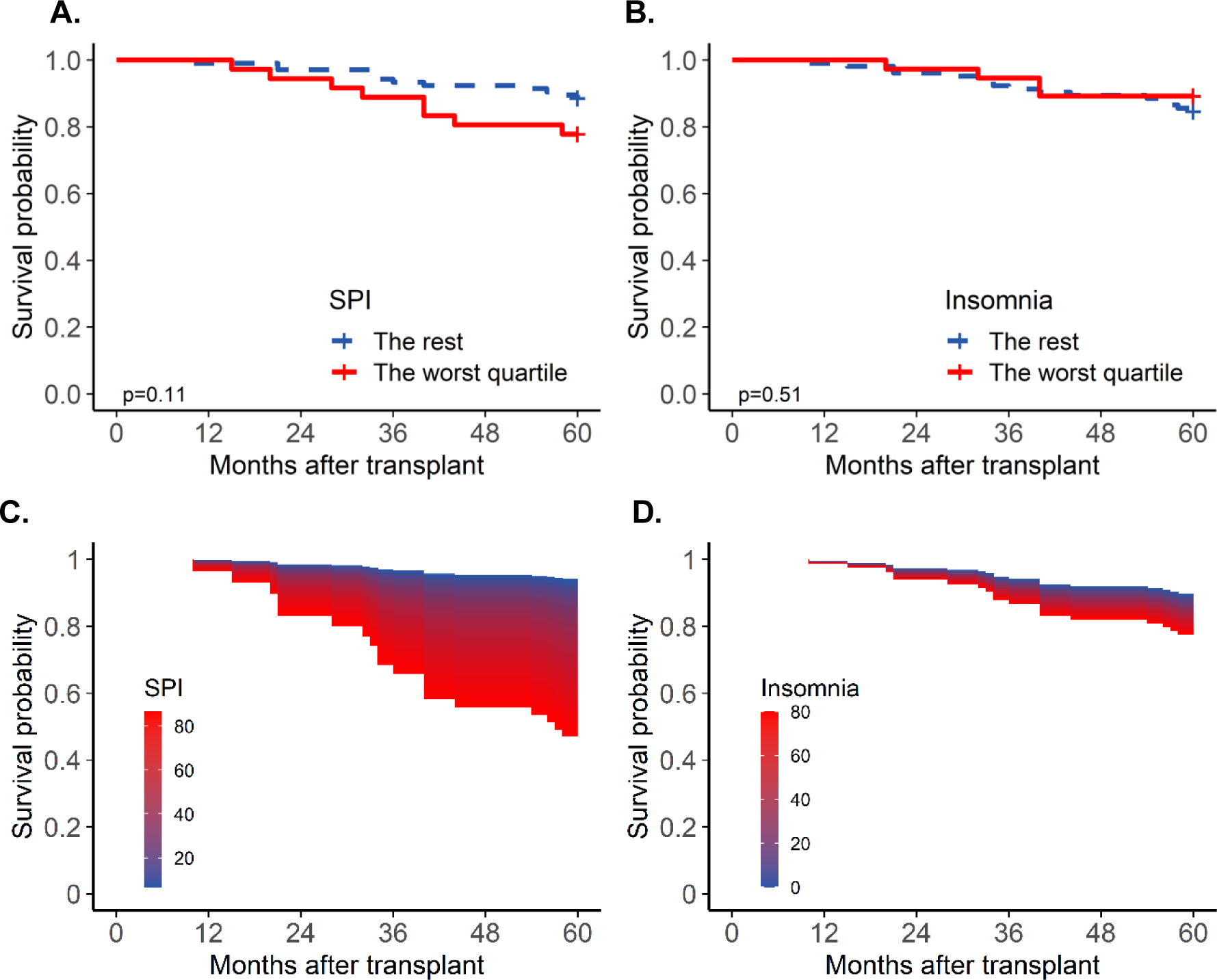
Kaplan Meier estimate of the association of disturbed sleep by MOS-Sleep Problems Index (panel A) and insomnia-specific subscale (panel B) defined as categorical variables on time to death. Survival Area Plot illustrating the association of disturbed sleep by MOS-Sleep Problems Index (panel C) and insomnia-specific subscale (panel D) defined as continuous variables on time to death.

**Table 4.** Association between sleep problem index and insomnia subscale with CLAD and death adjusting for age, sex, and transplant indication.

|  |  | **Time to CLAD**<br>**Hazards Ratio** | **Time to Death**<br>**Hazards Ratio** |
| --- | --- | --- | --- |
| **Sleep Problem**<br>**Index (SPI)**<br>SPI MID = 8 | **Per MID worsening** | 1.14 (1.00, 1.30)<br>p = 0.04 | 1.29 (1.05, 1.58)<br>p = 0.01 |
|  | **Worst quartile** | 2.18 (1.22, 3.89)<br>p = 0.01 | 2.12 (0.84, 5.35)<br>p = 0.11 |
| **Insomnia**<br>Insomnia MID = 12 | **Per MID worsening** | 1.21 (1.05, 1.39)<br>p = 0.01 | 1.16 (0.93, 1.44)<br>p = 0.20 |
|  | **Worst quartile** | 1.96 (1.09, 3.53)<br>p = 0.03 | 0.68 (0.22, 2.10)<br>p = 0.51 |

## Discussion

In this single-center study, we found that prolonged hospital stay after lung transplantation was associated with disturbed sleep and insomnia which, in turn, were associated with worse cognitive functioning, depressive symptoms and poorer HRQL. Disturbed sleep and insomnia were also associated with disability and frailty as well as increased risk of CLAD and mortality.

Our findings are consistent with evidence that emphasizes the critical role sleep plays in physical and mental health outcomes. They also add important insights into the relatively little that is currently known about the impact sleep has on patient-reported outcomes after lung transplantation. Among a small number of modest sized studies, poorer sleep after lung transplant has been associated with worse depressive symptoms and poorer scores on the general mental health component of the SF-36..^40–43^ An understanding of poor sleep on other outcomes, however, remains largely unknown. Outside of lung transplantation, poorer sleep is associated with cognitive dysfunction and risk of frailty.^16,18,44–46^ For example, experimental and epidemiologic data supports links between poor sleep and impaired cognition, including difficulties in executive functioning, memory, and attention processes.^17^ Notably, cognitive dysfunction following major cardiothoracic surgery is common and, in lung transplant, is associated with increased risk of mortality.^26,47–49^ Sleep disturbance during this important recovery period may contribute to future cognitive outcomes. Further, a pooled analysis supports short and excessively long sleep duration, as well as a longer sleep onset latency as risk factors for frailty.^18^

Our study has several notable strengths. First, it is one of the largest studies of sleep and patient reported outcomes in lung transplant. A recent scoping literature review of sleep quality following transplant identified just nine research studies with sample sizes ranging from 20 to 219 participants. Our study is the second largest to date and the first to link measures of sleep quality to outcomes beyond mental health. We newly identified associations between sleep and physical disability, frailty, CLAD, and even death after lung transplant, raising the possibility that sleep disturbance may be an important and modifiable behavioral pathway to improve quality life and clinical outcomes among lung transplant recipients.

Despite these strengths, it is important to highlight that our study design precludes causal inference testing or understanding mechanisms of association. For example, our measures of sleep were collected concurrent to other PROs. Thus, it remains unclear whether disturbed sleep caused depressive symptoms, poorer HRQL, and other PROs or whether the reverse may be true. Understanding the directionality of these relationships are key to further efforts including designing interventions aimed at improving patient-centered outcomes. Further, while the associations between disturbed sleep and cognitive, psychological, and HRQL outcomes may be more direct, the associations with physical impairments including frailty and CLAD may be less-so. Behaviorally-driven factors such as impaired motivation to exercise regularly and to consume a balanced diet could drive disability and frailty whereas depression and cognitive impairment could impact medication adherence, thereby increasing risk of CLAD. It is also plausible that sleep’s essential role in immune system dysregulation from disturbed sleep via enhanced systemic inflammation^20,50^ and accelerated biological aging^19,51^ *could* identify biological links between sleep and disability, frailty and CLAD. While plausible, these potential explanations should be considered speculative until more definitive research is performed.

There are additional limitations to bear in mind. This cohort was not originally designed to study the impact of sleep disturbances on outcomes after lung transplantation. Our convenience sampling strategy yielded a group of participants whose sleep was queried over a range of early post-operative years potentially limiting our ability to identify important peri- and post-operative predictors of poor sleep after transplant. We also lack information on how sleep disturbances change over the early post-operative period. In prior work by our team and by others, other factors after transplant improve for many in the first post-operative year. Whether *persistently* poor sleep is differentially associated with clinical outcomes than sleep that worsens or improves over time is unknown. Answers to this unknown are important when considering when to screen for sleep after transplant and when designing potential interventions. We also did not perform objective tests of sleep, which precluded us from determining which type of sleep impairment drove our findings. This has critical treatment implications as cognitive behavioral and pharmacologic interventions used to treat problems like insomnia are less effective for sleep disordered breathing conditions. While our insomnia subscale has face validity, we did not use a validated insomnia-specific measure. Thus, our findings related to insomnia should be interpreted with caution. Finally, although this cohort represents one of the largest studies of sleep after lung transplantation, it, nevertheless, is relatively modest in size.

Future efforts to study sleep longitudinally in the peri- and early post-operative period may help shed light on the causal role of sleep in lung transplant recovery. If our findings are confirmed, treatments for disturbed sleep attributable to disorders like insomnia and obstructive sleep apnea are effective, relatively easy to implement, and could be incorporated into post-transplant care both in person and remotely. Further, if our findings linking disturbed sleep with CLAD and mortality are confirmed, efforts to disambiguate their behavioral or immunological underpinnings could shed new insights into important outcomes in lung transplantation.

## Data Availability

All data produced in the present study are available upon reasonable request to the authors

## Acknowledgements

The authors are deeply appreciative of the time our patient participants provided to support this research and members of the UCSF clinical Advanced Lung Disease and Transplant program

