## Supplemental Material for "Disturbed sleep after lung transplantation is associated with worse patient-reported outcomes and chronic lung allograft dysfunction"

**Appendix Table 1.**

| | Continuous scale<br>SPI MID = 8 Insomnia MID = 12 | | Worst Quartile | |
|---|---|---|---|---|
| | **Sleep Problem Index (SPI)** | | | |
| **Predictor** | **Parameter estimate (95% CI)** | **p-value** | **Odds ratio (95% CI)** | **p-value** |
| Severe PGD | 3.80 (-2.66, 10.27) | 0.25 | 0.97 (0.39, 2.45) | 0.95 |
| Length of hospital stay >30 days | 12.18 (3.38, 20.99) | 0.01 | 7.09 (2.10, 23.95) | <0.01 |
| Delirium | -1.14 (-10.59, 8.30) | 0.81 | 2.40 (0.69, 8.38) | 0.17 |
| | **Insomnia** | | | |
| Severe PGD | 1.51 (-7.63, 10.64) | 0.74 | 0.72 (0.27, 1.92) | 0.51 |
| Length of hospital stay >30 days | 14.56 (2.08, 27.04) | 0.02 | 1.69 (0.47, 6.04) | 0.42 |
| Delirium | -0.27 (-13.28, 12.74) | 0.97 | 0.67 (0.13, 3.40) | 0.63 |

**Appendix Table 2.**

| | | **Time from survey to CLAD (Hazards Ratio)** | **Time from survey to Death (Hazards Ratio)** |
|---|---|---|---|
| **Sleep Problem Index (SPI)** MID = 8 | **Per MID worsening** | 1.09 (0.93, 1.27)<br>p = 0.29 | 1.27 (1.02, 1.57)<br>p = 0.03 |
| | **Worst quartile** | 1.76 (0.90, 3.45)<br>p = 0.10 | 2.06 (0.82, 5.18)<br>p= 0.12 |
| **Insomnia** MID = 12 | **Per MID worsening** | 1.15 (0.98, 1.36)<br>p = 0.09 | 1.10 (0.88, 1.38)<br>p = 0.41 |
| | **Worst quartile** | 1.73 (0.87, 3.47)<br>p = 0.12 | 0.64 (0.21, 1.96)<br>p = 0.43 |
